# Cluster Analysis to Find Temporal Physical Activity Patterns Among US Adults

**DOI:** 10.1101/2023.01.23.23284777

**Authors:** Jiaqi Guo, Marah M. Aqeel, Luotao Lin, Saul B. Gelfand, Heather A. Eicher-Miller, Anindya Bhadra, Erin Hennessy, Elizabeth A. Richards, Edward J. Delp

## Abstract

Physical activity (PA) is known to be a risk factor for obesity and chronic diseases such as diabetes and metabolic syndrome. Few attempts have been made to pattern the time of physical activity while incorporating intensity and duration in order to determine the relationship of this multi-faceted behavior with health. In this paper, we explore a distance-based approach for clustering daily physical activity time series to estimate temporal physical activity patterns among U.S. adults (ages 20-65) from the National Health and Nutrition Examination Survey 2003-2006 (NHANES). A number of distance measures and distance-based clustering methods were investigated and compared using various metrics. These metrics include the Silhouette and the Dunn Index (internal criteria), and the associations of the clusters with health status indicators (external criteria). Our experiments indicate that using a distance-based cluster analysis approach to estimate temporal physical activity patterns through the day, has the potential to describe the complexity of behavior rather than characterizing physical activity patterns solely by sums or labels of maximum activity levels.

## I. Introduction

Physical activity (PA), the movement produced by the contraction of skeletal muscle causing energy expenditure above a basal level, is a complex human behavior providing many health benefits regardless of age, gender, race, fitness level, or current life stage [2]. Physical activity is a risk factor for obesity and chronic diseases such as diabetes and metabolic syndrome [34], [35], [44], [47], [53]. Studies that have investigated patterning physical activity lag behind other health behaviors, such as diet, both conceptually and methodologically. To date, work has mainly focused on physical activity patterns of varying intensities and how they link to health [41], [43], [46], [48], [49]. Less is known about the patterns of physical activity behaviors over continuous time and their associations to health status indicators such as body-mass-index (BMI) and waist circumference. Recent studies provide evidence that the time of physical activity may be related to health [8], [13], [55], [56], and a few heuristic concepts of temporal physical activity patterns have been introduced to define the chronological succession of physical activities [26], [46]. Other studies have attempted to include the time of the physical activity by using summary estimates such as the percentage of morning activity to daily total activity [13] or the time of the exercise session [8]. These studies provided early developments of temporal physical activity patterns, but relied heavily on experts’ knowledge and pre-defined standards. Using data-driven methods such as cluster analysis for the incorporation of time, intensity, and other factors of physical activity is rarely considered. The computational limitations incurred by using complex distance measures have further hindered the analysis of “minute-level” accelerometer-collected data from physical activity studies.

In this paper, we define the creation of Temporal Physical Activity Patterns (TPAP) as the partitioning of daily physical activity time series data into mutually exclusive clusters using data-driven approaches that incorporate the time of activities as well as other factors such as intensity and duration. Each cluster represents a specific temporal pattern of physical activity. This paper describes a distance-based cluster analysis approach for estimating temporal physical activity patterns using the National Health and Nutrition Examination Survey 2003-2006 (NHANES) [3]. The purpose of our work is to investigate the creation of the TPAPs including intensity, time, and duration, and to link the TPAPs to health.

The NHANES physical activity dataset (1999 participants after exclusion) [3] was collected using uni-axial accelerometers that measures vertical acceleration in units known as “physical activity count (PAC)”. Participants’ physical activity data correspond to one-dimensional time series of length 1440 minutes (60 × 24). The samples of the time series denote the summed PACs in each one-minute epoch. Since daily physical activity time series are subject to nonlinear warpings in time (e.g., walking to work 10 minutes earlier, jogging for 45 minutes instead of 30), the samples from two time series need to be aligned before being compared. We choose Dynamic Time Warping (DTW) [42] to align the samples to be able to compare the physical activity times series. To prevent potential pathological warpings (e.g., aligning activities in the morning to activities in the evening) and take into consideration the temporal nature of the physical activity, the Sakoe-Chiba Band [42] is introduced as a global constraint which limits the maximum temporal difference between matched activities.

The DTW distance is combined with three distance-based clustering methods that are commonly used in time series clustering, namely kernel k-means [40], spectral clustering [30], and hierarchical agglomerative clustering [37]. The distance-based clustering methods require the computation of a distance matrix which consists of the pairwise DTW distances of all participants in the dataset. Since the DTW distances for different pairs of participants are computed independently, we use Graphics Processing Unit (GPU) and CUDA parallel computing platform [31] to accelerate the computation of the distance matrices. Our goal is to select the distance measures and clustering methods that generate clusters with distinctive physical activity characteristics and meaningful links to health. In this paper, we investigate three approaches for evaluating the clustering results: (1) internal criteria (the Silhouette Index and the Dunn Index [17]), (2) external criteria (associations between clusters and health status indicators determined by multivariate linear regression), and (3) cluster visualizations including mean trajectory and heat map.

Our major contributions in this paper can be summarized as follows:

- We investigate a distance-based cluster analysis approach for estimating temporal physical activity patterns in the NHANES dataset.
- We evaluate the clustering results through internal criteria, external criteria, and visualizations.
- We show through experiments that the temporal physical activity patterns are derived based on the integration of time and intensity of physical activity, and could meaningfully link to health.

## II. Related Work

### A. Clustering Methods for Physical Activity Data

Here we review previous methods for clustering physical activity data. The physical activity data used in this paper are time series representing the intensity of physical activity in a day. While searching for related literature, only a small number of works used the same daily physical activity time series to study the association to health. Therefore, we also include in the review some works that used different type of physical activity data (not time series), but focused on the association between physical activity and health. These methods for clustering physical activity data can be categorized into two types: 1) using pre-defined grouping standards (summary metric); and 2) using data-driven methods such as k-means and hierarchical clustering.

Pre-defined grouping standards or summary metrics are usually designed by domain experts to separate the participants into clusters [8], [13], [26], [43], [46], [49]. In [8], Alizadeh et al. divided 48 overweight females into morning and evening groups based on the time of exercise to study the effect on appetite and anthropometric indices. Chomistek et al. [13] clustered the participants into quartiles based on the ratio of activities before noon, and studied the clusters’ association to body mass index (BMI). Lindgren et al. [26] summarized accelerometer data into 3 pre-defined levels: sedentary, low, and medium-to-vigorous, and used regression models to find links between physical activity levels and socioeconomic status. In general, using pre-defined grouping standards has the flexibility in analyzing desired physical activity features. However, the grouping standards often omit detailed information from original, minute-level physical activity data. In addition, designing the grouping standards requires domain expert knowledge, which potentially limits the quality of physical activity patterns [54]. There is little evidence of what are the “correct” grouping standards, and the appropriate intensity, duration, and time of physical activity throughout a day [2].

Data-driven methods have also been investigated to derive unbiased interpretation of physical activity patterns [7], [39], [41], [52]. The data-driven methods followed two general directions. The first one is the model-based approach which constructs probabilistic models for fitting physical activity data. In [41], Metzger et al. first determined total minutes of moderate-to-vigorous physical activity (MVPA), vigorous physical activity (VPA), and minutes of MVPA that occurs in bouts of 10 minutes for each participant. These intensity features were used to design a latent class analysis (LCA) model to cluster the NHANES dataset and estimate physical activity patterns associated with the clusters. The second direction is the distance-based approach using features from descriptive statistics or important motifs [7], [39], [52]. Rovniak et al. [39] examined the duration of moderate-to-vigorous physical activity (MVPA) from four different life domains: leisure, occupation, transport, and home. The authors then used the Euclidean distance and hierarchical agglomerative clustering to group the participants and study psychosocial and built-environment differences in U.S. adults. These datadriven methods mainly focused on varying physical activity intensities, while the time of activity (temporal information) was often omitted.

In this paper, we also explore a distance-based approach. In contrast, the distances we use are computed using DTW based on the daily physical activity time series instead of intensity features so that the time of the activity is considered. Dynamic Time Warping (DTW) and similar elastic distances [28] have been widely used in time series clustering/classification [29], [45], [57]. There have also been some works using DTW to analyze daily physical activity time series. In [54], the authors used Low Rank and Sparse matrix decomposition (LRS) to decompose a daily physical activity time series dataset (HealthyTogether [11]). They used DTW and hierarchical agglomerative clustering on the low rank and sparse components separately, and generated the final clusters through cross-product. However, global constraints for DTW and alternative clustering methods were not explored in [54], and the evaluation of clustering results is limited to internal criteria and visualizations. The relationship between physical activity patterns and health was not studied in [54] due to the lack of anthropometric and health-indicator data.

### B. NHANES 2003-2006 for Physical Activity Studies

The NHANES 2003-2006 [3] is one of the only publicly-available, nationally representative datasets to capture physical activity through accelerometry devices [4], [5]. Here we review how the previous works used the NHANES 2003-2006 for physical activity research. Among the literature we reviewed, only a few used clustering methods to separate the participants in the NHANES 2003-2006 dataset and studied their association to health [14], [41], but the physical activity data were simplified to represent only the intensity, while the time of activities was often omitted. In some studies [10], [15], [27], [32] the participants were clustered based on their anthropometric or health-indicator data (e.g., age, gender, BMI, etc.), then the authors studied the association between the clusters and physical activity. There are also some works which used the NHANES physical activity data to perform regression analysis against health status indicator [20], [33]. To our knowledge, few previous works used the NHANES 2003-2006 dataset to perform clustering analysis based on the minute-level, accelerometer-collected physical activity time series.

## III. Our Proposed Approach

### A. DTW Distance Measure

Let **X** = [*x*_1_, *x*_2_, …, *x*_*M*_] and **Y** = [*y*_1_, *y*_2_, …, *y*_*M*_] be two one-dimensional time series of the same length *M*. Let **X**[*k* : *l*] = [*x*_*k*_, *x*_*k*+1_, …, *x*_*l*_] be the sub time series of **X** (1 ≤ *k* ≤ *l* ≤ *M*). In this paper, these time series represent the daily activity routines of different participants. The samples *x*_*i*_ and *y*_*j*_ are the Physical Activity Counts (PACs) collected using uniaxial accelerometry devices. Their time indices *i* and *j* denote the time (minute) when *x*_*i*_ and *y*_*j*_ are collected. Detailed description of the NHANES physical activity dataset can be found in section IV-A.

The first step in estimating temporal physical activity patterns is to find a proper distance measure for comparing the physical activity time series. Two physical activity time series **X** and **Y** may not be aligned temporally. This is due to the fact that different participants have different daily activity routines. The Euclidean distance between **X** and **Y** is defined as:

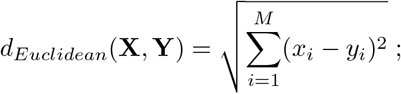

Using the Euclidean distance, a small temporal shift in **X** or **Y** could result in a large increase in the distance, which is unsuitable for physical activity analysis. The Dynamic Time Warping (DTW) [42] distance provides a way of temporally aligning two time series such that the distance measure is not sensitive to temporal misalignment. The alignment of time series **X** and **Y** determined by DTW is denoted as the optimal warping path *P*. The optimal warping path *P* is a set of time index pairs that describes how the samples of **X** map into the samples of **Y** (e.g., if the *i*^*th*^ sample of **X** is mapped to the *j*^*th*^ sample of **Y**, the time index pair (*i, j*) will be included in the warping path). The DTW distance between **X** and **Y** is defined as

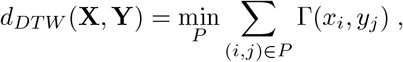

where Γ : ℝ × ℝ → ℝ^+^ is a local distance function. Details about finding the optimal warping path can be found in [42]. In this paper, we choose Γ(*x*_*i*_, *y*_*j*_) = (*x*_*i*_ − *y*_*j*_)^2^ to be the local distance function.

Fig. 1 gives an example of how two physical activity time series from the NHANES dataset are aligned by the Euclidean distance and the DTW distance respectively. Here, y-axis is the physical activity count (PAC) which represents the intensity of activities (see Section IV-A for more detail). With the Euclidean distance, the intensive activities of one participant are often aligned to the sedentary periods of another. This exaggerates the actual difference between the two participants’ activity patterns. The DTW distance aligns the two time series such that the intensive activities from one participants are compared to the intensive activities of another. In this paper, to prevent potential pathological alignments (e.g., aligning morning activities to evening activities), the Sakoe-Chiba band [42] is used as a constraint on the warping path of DTW. The Sakoe-Chiba band limits the maximum difference between the time indices of aligned samples, i.e., it limits (*i, j*) ∈ *P* by |*i* − *j*| ≤ *T*, where *P* is the warping path, and *T* is the parameter for the Sakoe-Chiba bandwidth. We denote constrained DTW with the Sakoe-Chiba Band as CDTW.

**Fig. 1.**
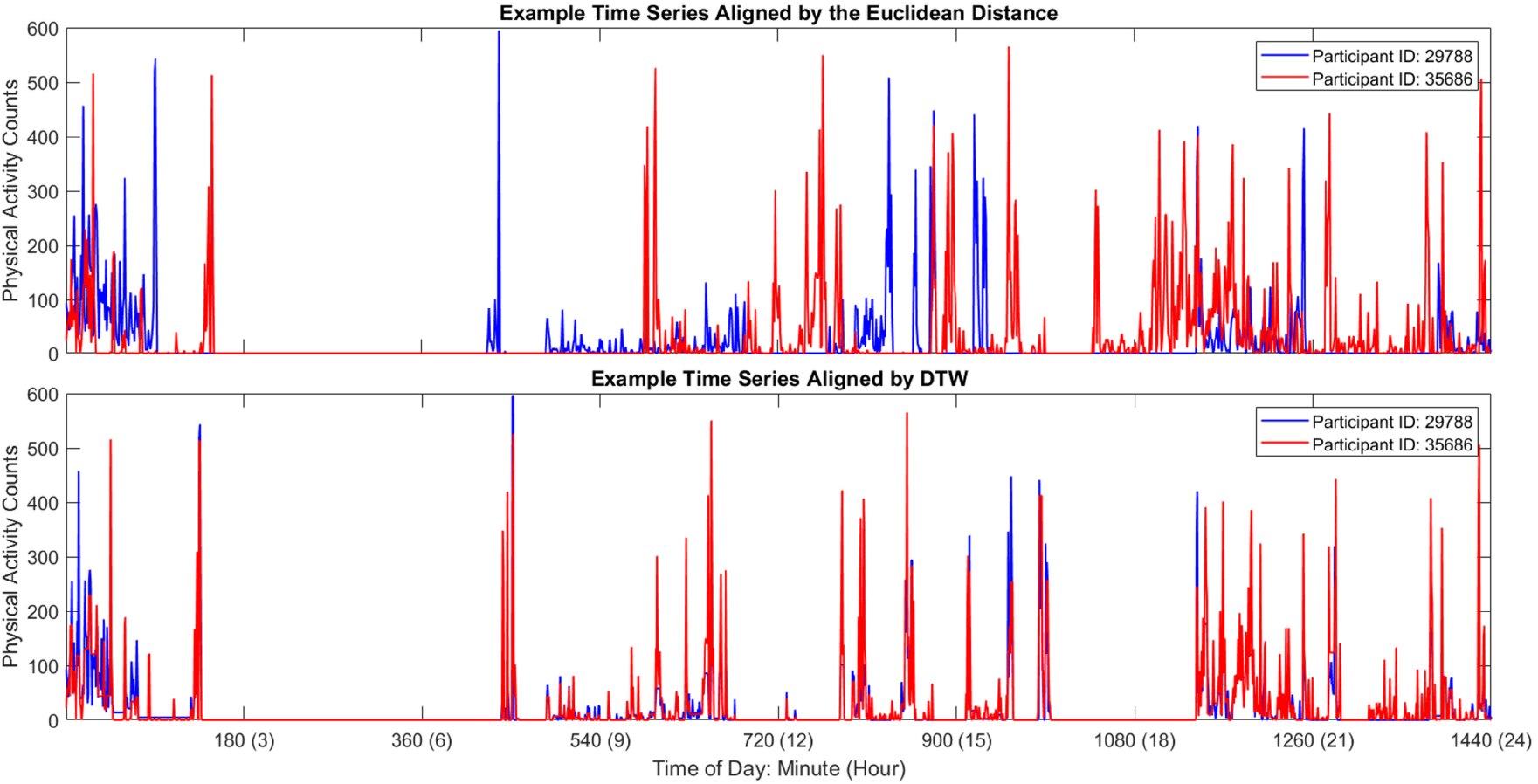
Temporal alignment of two physical activity time series from the NHANES dataset. Top: The original time series before time warping. The Euclidean distance compares the samples with the same time index, and exaggerates the actual difference. Bottom: The DTW distance aligns the samples to compensate the temporal shifts.

### B. Clustering Methods

The second step of estimating temporal physical activity patterns is to group the time series based on the selected distance measure. Several distance-based clustering methods are examined in the Experimental Section IV. Based on our experimental results we focus on the most successful approaches. These include kernel k-means [40] and kernel hierarchical agglomerative clustering with Ward’s Linkage [24].

#### 1) Kernel K-Means

Kernel k-means has been used to cluster data points which are not linearly separable. First, the time series are projected from the input space *V* onto a hyperspace *W* using a projection function *ϕ* : *V* → *W*. Next, the projected points in the hyperspace are partitioned using the standard k-means clustering method. The inner product of projected points in hyperspace *W* can be computed through the corresponding kernel function *k* : *V* × *V* → R which satisfies

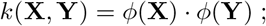

Let 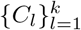 denote a partition of the dataset into *k* clusters, and let 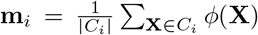 be the “center” of the *i*^*th*^ cluster in the hyperspace. Kernel k-means finds the partition by minimizing an objective function defined using the total sum of squared errors

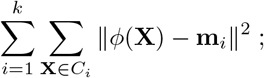

Furthermore, the squared distance from point **X** to the center of the *i*^*th*^ cluster **m**_*i*_ can be written as

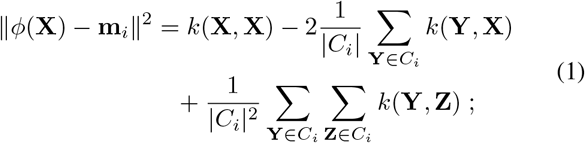

Equation (1) shows that as long as the corresponding kernel function *k* is known, the projection function *ϕ* is not needed in computing distances between cluster centers and data points in the hyperspace. Hence a kernel function matrix **K** consisting of the pairwise kernel function values of all data points is sufficient to perform kernel k-means clustering without the original data points.

#### 2) Kernel Hierarchical Agglomerative Clustering

Hierarchical Agglomerative Clustering (HAC) is frequently used in physical activity cluster analysis [39], [54]. The HAC starts by treating each data point as a cluster, and then successively merges two nearest clusters by minimizing a measure of pairwise cluster distance, until there is a single cluster. A dendrogram [37] is formed during the agglomeration process, and the desired number of clusters can be drawn from the dendrogram.

There are several different ways of defining the distance between clusters, which are referred to as linkage methods. Some commonly used linkages are based on the spatial distances between data points, such as Single, Complete, and Average [37]. To use kernel functions on these linkages, one can use agglomeration based on the distances in hyperspace [24]:

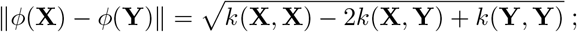

However, using these linkages (Single, Complete, Average) to the CDTW distance matrices leads to a serious chaining effect [37], where the data points sequentially get merged into a single cluster. From our experimental observations, the generated clusters are greatly unbalanced in size, with the largest cluster taking up to 95% of the entire dataset. Such issues limit the fidelity of the external evaluation of the clusters. The unbalanced clusters also tend to emphasize extreme patterns that only represent a relatively small number of participants, and do not support our goal of defining population physical activity patterns.

In contrast, kernel Ward’s method [24] is a linkage method that aims to minimize cluster variance. The variance of a cluster *C*_*i*_ is defined using the sum of squared errors

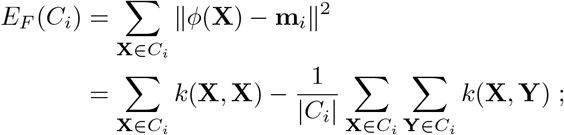

To determine whether clusters *C*_*i*_ and *C*_*j*_ are next to be merged, the distance between them is defined as

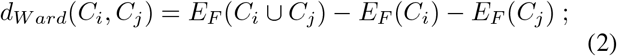

The distance between the merged cluster *C*_*i*_ ∪ *C*_*j*_ and another cluster *C*_*q*_ is then updated by

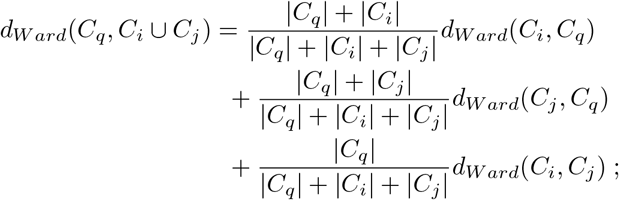

Notice that kernel Wards’s method and kernel k-means both attempt to minimize the cluster variance in hyperspace. In our experiments (Section IV-B), these two methods produce more balanced clusters, suggesting that for the physical activity data the total sum of squared errors is an appropriate objective function.

#### 3) Convert CDTW into Kernel Function

In this paper, we used the Gaussian Dynamic Time Warping Kernel [9] to convert the CDTW distances into kernel function values for kernel k-means and kernel hierarchical agglomerative clustering:

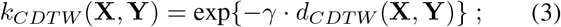

The Gaussian DTW Kernel is straightforward and easy to implement. In this paper, we further exploited the parallel structure of GPUs to accelerate the computation of CDTW distance matrices.

## IV. Experiment and Evaluation

### A. Dataset

The NHANES is designed to assess the health and nutritional status of children and adults in the United States [3]– [5]. In order to represent the non-institutionalized civilian U.S. population, a multi-stage probability sampling design is used to recruit participants for NHANES. To date, the NHANES is one of the only publicly-available, nationally representative datasets to capture physical activity through accelerometry.

The physical activity data were collected as follows. Participants wore an ActiGraph AM-7164 (Formerly the CSA/MTI AM-7164; ActiGraph, LLC, Fort Waltion Beach, FL) on the right hip. The Actigraph AM-7164 uses a cantilever beam sensor for measuring vertical acceleration in units known as “physical activity count (PAC)” at a sampling frequency of 10 Hz. The original data from the accelerometer were filtered using analog methods, and typically calibrated in a laboratory to achieve linear regression associations between the “counts” and a measured physiologic variable [21]. In the NHANES 2003-2006 Examination, orginal PACs (10 Hz) were further summed over each minute epoch [3]. All participants were asked to wear the devices on for 7 consecutive days. Only non-pregnant participants who were 20-65 years without missing anthropometric and laboratory data are included in the experiments. Due to compliance and other factors, there are a large number of participants who do not have a full 7-day record. To maximize the number of participants involved in our analysis, we included anyone with at least one weekday of valid accelerometer data. There are 1999 targeted participants after exclusions, and one valid weekday of each participant is randomly selected to form the physical activity dataset. Note that the physical activity data of each participant corresponds to a one-dimensional time series:

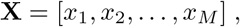

where *x*_*i*_ is the physical activity counts (PACs) summed over the *i*^*th*^ minute in a day, and *M* = 1440 is the number of minutes in a day (24-hour period). Two example time series can be found in Figure 1.

### B. Cluster Evaluation

Participants were clustered using methodologies discussed in the previous section. For parameter selection, the Gaussian DTW Kernel parameter *γ* in Equation 3 is fixed to be half of the average of all distances in the CDTW distance matrix. With different values of Sakoe-Chiba bandwidth *T*, the CDTW distance would have varying emphasis on the time difference of physical activity over intensity difference. In this paper, we investigated 12 values of *T* ranging from 60 to 720 (minute) in step of 60. We did not keep increasing the bandwidth to the maximum (1440) because the CDTW distances of most pairs of participant converge to DTW before bandwidth 720. Keeping increasing the bandwidth after *T >* 720 has little influence on the generated clusters. CDTW with different bandwidths can be treated as different distance measures. We combined each distance measure with all three clustering methods (kernel k-means, spectral clustering, and kernel hierarchical method) to have a total of 36 different combinations. For the number of clusters, *k*, we explored four values *k* ∈ {3, 4, 5, 6} for each combination of distance measure and clustering method. We conducted 144 experiments on the NHANES physical activity dataset (12 distance measures × 3 clustering methods × 4 numbers of clusters). From all the combinations of distance measures and clustering methods we explored, we wish to select the ones that could generate clusters with both distinctive physical activity characteristics and meaningful links to health.

As our proposed approach is essentially a time series clustering method, the evaluation of the clustering results is not easy due to the absence of ground truth. In a review of time series clustering studies [6], three possible approaches for evaluating clustering results were described: internal criteria, external criteria, and visualizations. In this paper, all three evaluation approaches were used. For robust post hoc tests, we also take into consideration the sample sizes of the clusters.

#### 1) Sample Sizes of Clusters

As suggested by the NHANES analytic guidelines [1], a cluster size of less than 30 is considered insufficient for inferential analysis based on the normal approximation. Therefore, we disregard the unbalanced clustering results whose smallest cluster has less than 30 participants. From our experiments, the unbalanced clusters appeared most often in clustering results derived by spectral clustering and kernel hierarchical clustering with Single, Complete, or Average linkage methods. Of all these results, the largest clusters always take up the majority of the participants in the dataset (over 90%), leaving few participants in the smaller clusters (in most cases less than 10). Following the instructions of the NHANES analytic guidelines [1], we disregard the results derived by spectral clustering and kernel HAC with Single, Complete, and Average linkage, and focus on kernel k-means and kernel HAC with Ward’s linkage.

#### 2) Internal Criteria

The internal criteria are similar to the objective function in kernel k-means, and formalize the goal to reach higher intra-cluster similarity and lower inter-cluster similarity [6]. Since the internal criteria are computed based on the PA data, they evaluate whether the clusters from the same result have distinctive physical activity characteristics. Two internal criteria that are commonly used in cluster analysis were adopted in this paper, namely the Silhouette Index [38] and the Dunn Index [17]. For both criteria, higher values indicate better clustering quality. When computing the internal criteria, we always follow the same distance measure that was used to generate the clusters.

When ground truth is not available, the number of clusters is commonly determined through internal evaluations [41], [54]. Table I lists the Silhouette and the Dunn Index for different number of clusters *k* while fixing clustering method to kernel k-means. For both internal criteria and most combinations of distance measures and clustering methods, the smaller number of clusters generally achieves better internal criteria. Therefore, we choose the number of clusters to be *k* = 3 in the following discussions. In some related studies, the internal criteria were also used to evaluate distance measures and clustering methods. However, it can be shown from our experiments (Table II) that the Silhouette Index and the Dunn Index may support different clustering methods at certain CDTW bandwidths (e.g. when *T* = 540). If we fix kernel k-means as clustering method and compare different distance measures, we can find that while the Silhouette Index favored CDTW with smaller bandwidth, larger CDTW bandwidths generally achieved better Dunn Index. It is worth mentioning that different internal criteria have their own ways of defining inter- and intracluster difference [17], and it remains arguable whether internal criteria are valid for comparing distance measures. Therefore, we introduce the external criteria in the following section to help the evaluation of distance measures and clustering methods.

**TABLE 1.** The Silhouette/Dunn scores for different number of clusters *k* while fixing clustering method to kernel k-means.

| $T^a$ | $k = 3$ | $k = 4$ | $k = 5$ | $k = 6$ |
| --- | --- | --- | --- | --- |
| 60 | 0.257/0.062 | 0.167/0.054 | 0.088/0.045 | 0.069/0.025 |
| 120 | 0.214/0.062 | 0.151/0.053 | 0.059/0.035 | 0.053/0.035 |
| 180 | 0.201/0.069 | 0.149/0.065 | 0.085/0.051 | 0.057/0.053 |
| 240 | 0.138/0.067 | 0.148/0.070 | 0.055/0.057 | 0.070/0.057 |
| 300 | 0.145/0.076 | 0.057/0.068 | 0.056/0.064 | 0.054/0.063 |
| 360 | 0.147/0.078 | 0.057/0.071 | 0.056/0.068 | 0.057/0.063 |
| 420 | 0.148/0.081 | 0.056/0.073 | 0.060/0.073 | -0.002/0.046 |
| 480 | 0.150/0.083 | 0.058/0.074 | 0.064/0.074 | -0.001/0.047 |
| 540 | 0.148/0.084 | 0.059/0.075 | 0.065/0.074 | -0.001/0.048 |
| 600 | 0.147/0.084 | 0.059/0.074 | 0.065/0.075 | 0.001/0.048 |
| 660 | 0.152/0.084 | 0.060/0.074 | 0.066/0.075 | 0.002/0.048 |
| 720 | 0.148/0.085 | 0.061/0.074 | 0.066/0.075 | 0.003/0.048 |

<sup>a</sup> The Bandwidth of CDTW.

**TABLE 2.** The Silhouette Index, the Dunn Index (internal), and the number of total significant-difference pairs (external) for clustering results derived from different combinations of distance measures (rows) and clustering methods (columns) while the number of clusters was set to 3.

| $T^a$ | KKM<sup>b</sup> | | | KHAC-Ward<sup>c</sup> | | |
| --- | --- | --- | --- | --- | --- | --- |
|  | S<sup>d</sup> | D<sup>e</sup> | SDP<sup>f</sup> | S | D | SDP |
| 60 | 0.257 | 0.062 | 8 | 0.221 | 0.053 | 8 |
| 120 | 0.214 | 0.062 | 9 | 0.065 | 0.035 | 8 |
| 180 | 0.201 | 0.069 | 11 | 0.253 | 0.067 | 10 |
| 240 | 0.138 | 0.067 | 9 | 0.057 | 0.068 | 8 |
| 300 | 0.145 | 0.076 | 12 | -0.054 | 0.044 | 7 |
| 360 | 0.147 | 0.078 | 12 | 0.207 | 0.075 | 9 |
| 420 | 0.148 | 0.081 | 14 | -0.004 | 0.038 | 7 |
| 480 | 0.150 | 0.083 | 13 | 0.038 | 0.067 | 8 |
| 540 | 0.148 | 0.084 | 15 | 0.208 | 0.078 | 12 |
| 600 | 0.147 | 0.084 | 15 | 0.041 | 0.051 | 7 |
| 660 | 0.152 | 0.084 | 13 | 0.312 | 0.090 | 10 |
| 720 | 0.148 | 0.085 | 14 | 0.281 | 0.068 | 7 |

<sup>a</sup> The Bandwidth of CDTW; <sup>b</sup> Kernel K-Means; <sup>c</sup> Kernel HAC with Ward's Linkage; <sup>d</sup> the Silhouette Index; <sup>e</sup> the Dunn Index; <sup>f</sup> Number of Significant Difference Pairs.

#### 3) External Criteria

In general, the external criteria such as the Rand Index [36] evaluate clustering results based on ground truth. Unfortunately, participants in the NHANES physical activity dataset are not labeled. Instead, each participant has several health status indicators such as body mass index (BMI) and blood pressure. In this paper, we use these indicators to define an external score which represents the clusters’ association to health.

Twelve health status indicators were chosen for their previously studied associations with physical activity [12], [16], [50], including BMI, waist circumference (WC), fasting plasma glucose, triglycerides, hemoglobin A1c, total cholesterol, high-density lipoprotein (HDL-C), systolic blood pressure (SBP), diastolic blood pressure, type 2 diabetes (T2DM), metabolic syndrome (MS), and obesity. For one clustering result and a specific health status indicator, we use the cluster label as explanatory variable and the health status indicator as dependent variable to perform Multivariate Linear Regression (MLR). We control for potential confounders (survey year, age group, sex, ethnicity/race, poverty-income ratio (PIR), BMI, and total activity count (PACs summed over one day)) and adjust for multiple comparisons to determine the association between the clusters (explanatory variable) and the health status indicator (dependent variable). Tukey-Kramer’s procedure [51] was used for adjusting the p-values. In this paper, we chose the 5% significance level (*α*). Two clusters are considered significantly different if their adjusted p-value of pairwise comparison is less than 0.05. SAS (version 9.4) was used to complete the MLR and multiple comparison analysis. After one clustering result’s associations to all 12 health status indicators are determined, the external score is computed as the total number of pairwise comparisons that show significant difference. With more pairs of comparisons showing significant difference, there is a stronger association between the clusters and health. For detailed data collection procedures of the selected health status indicators and confounders, we refer interested readers to the Anthropometric Assessment and Laboratory Tests section in [18], [19], [25].

From all the methodologies considered in this paper, kernel k-means combined with CDTW bandwidth of 540 and 600 both have the highest external score (15 significant-difference pairs among the 12 health status indicators). Since CDTW bandwidth 540 achieved better internal scores than bandwidth 600, we choose kernel k-means combined CDTW bandwidth 540 to generate the final temporal physical activity patterns. In the following section, we will use visualization tools to show the physical activity characteristics of the clusters generated using this combination. Here we do not attempt to adjust our subsequent inferential analysis for the selection of the clustering parameters. But this is reasonable in view of the fact that unadjusted confidence intervals are generally much closer to those which are obtained with post selection inference (for example with lasso, where such a framework exists [22]) than what would have been obtained with data splitting due to loss of efficiency.

#### 4) Cluster Visualization

Unlike the internal and external criteria, visualizations do not have a scalar value that reflects the quality of clustering results. Instead, visualizations intuitively illustrate the physical activity characteristics of different clusters, such as the overall physical activity intensity level and the most active time periods in a day. In this paper, we visualized the clustering results through their mean trajectories and heat maps. Due to page limitations, we mainly focused on the visualizations of the selected clustering result (kernel k-means coupled with CDTW bandwidth 540). Details about the two visualization methods are given bellow.

**Mean Trajectory** of a cluster **C** shows the average intensity at each time unit. It is defined as:

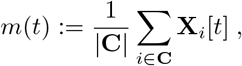

where **X**_*i*_ denotes the physical activity time series of a participant who gets partitioned to cluster **C. C** denotes the number of participants in cluster **C**. Time unit here is converted into hour level (*t* ∈ [0 : 23]) for better visualization, and **X**_*i*_[*t*] is the summed intensity over the *t*^*th*^ hour of the *i*^*th*^ participant.

**Heat Map** of a cluster **C** is a gray scale image which shows the distribution of non-zero physical activity counts (PACs) at each time unit. For cluster **C**, its physical activity heat map is defined as:

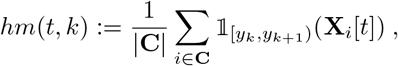

where *y*_*k*_ (*k* ∈ [0 : *K*] and *K* = 80) denotes an increasing sequence of physical activity intensity with *y*_*k*_ = *k* × Δ and Δ = 1500, which defines the intensity bins; 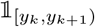 is the indicator function:

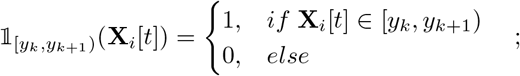

*y*_*K*_ (*K* = 80) is specially set to infinity to accommodate the extreme activity intensities.

Fig. 2 and Fig. 4 show the mean trajectories and heat maps of the clusters. Here, the constraint on time difference is relatively weak, and participants are separated based mainly on the intensity level. When bandwidth *T* reaches 540 (minute), the CDTW distances almost converged to DTW, and keeping increasing *T* has little influences on the clusters. Our observations show that the mean trajectories of clusters will not have significant changes for bandwidth larger than 540. From Fig. 2, all three clusters’ most active periods in a day are from 8 a.m. to 4 p.m., and participants become sedentary after 4 p.m. Cluster 2 is the most active of all three clusters, but has the smallest number of participants. Cluster 1 contains the largest number of participants and has moderate amount of activities. Cluster 3 also contains a large number of participants, but has the lowest activity level. Similar physical activity characteristics can be observed in their heat maps as well.

**Fig. 2.**
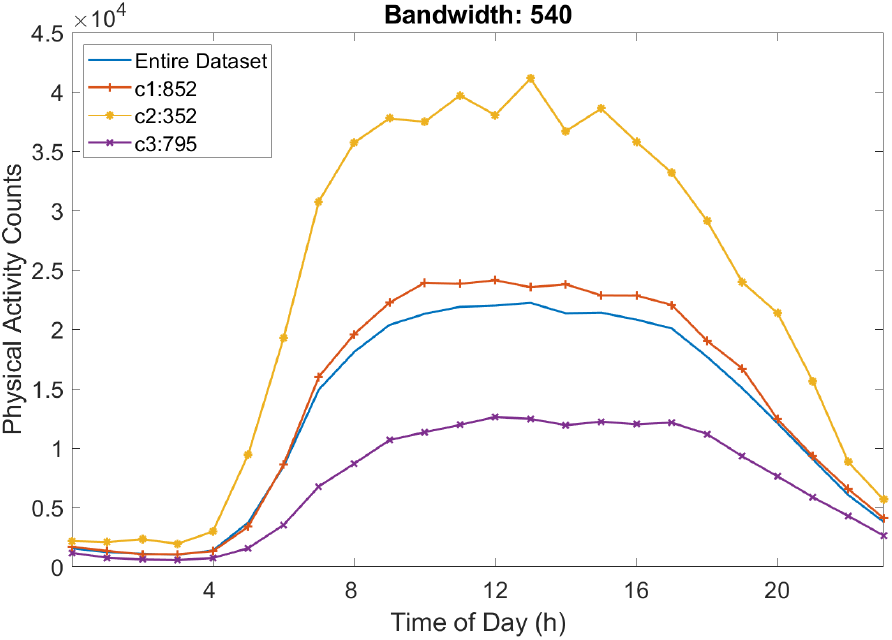
Mean trajectories of the three clusters obtained using kernel k-means and CDTW with *T* = 540.

As we decrease the bandwidth *T*, CDTW distance and clustering results focus more on participants’ temporality difference than intensity difference. Fig. 3 shows the mean trajectories of the clusters obtained using CDTW bandwidth 60 and kernel k-means. Comparing Fig. 3 to Fig. 2, the temporality difference between clusters in Fig. 3 becomes much more obvious. Here, Cluster 1 is the most sedentary cluster, and has a lower intensity level throughout the day; Cluster 2 and Cluster 3 both have more active physical activity patterns, but the most active period of Cluster 2 takes place between 8 a.m. and 12 a.m. in the morning while for Cluster 2 it happens between 4 p.m. and 8 p.m. in the late afternoon.

**Fig. 3.**
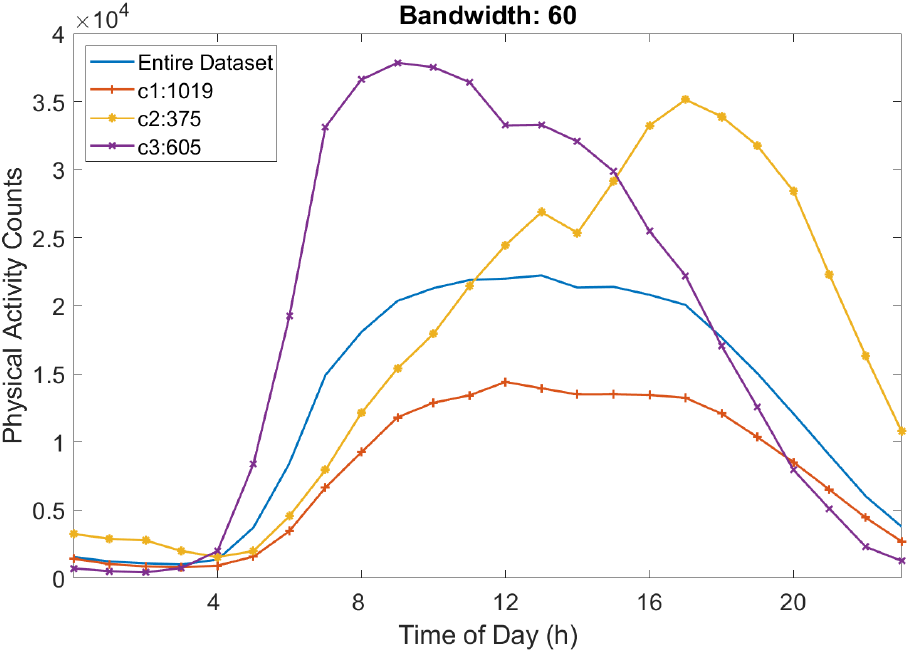
Mean trajectories of the three clusters obtained using kernel k-means and CDTW with *T* = 60.

**Fig. 4.**
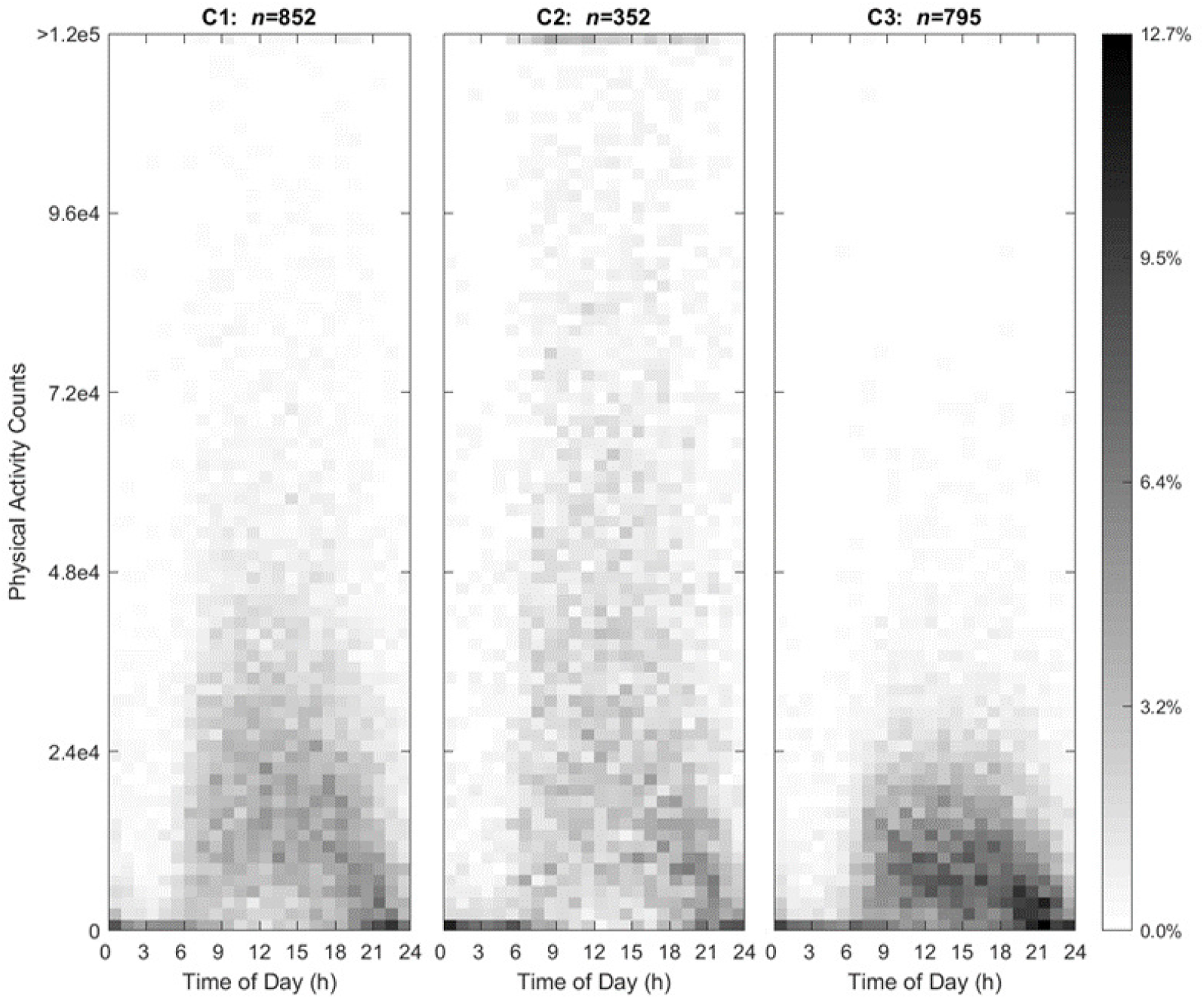
Heat Maps of the three clusters obtained using kernel k-means and CDTW with *T* = 540. From left to right: Cluster 1 to 3.

## Conclusion

In this paper, we describe a cluster analysis approach that combines distance measures and distance-based clustering method for estimating temporal physical activity patterns of U.S. adults ages 20-65 in the NHANES dataset. From our exploration on clustering methods, using cluster-variance based objective functions is likely to have a positive effect on producing more equal-sized clusters. Therefore, we mainly focused on kernel k-means and kernel HAC with Ward’s linkage in the evaluation section. We adopted both internal criteria and external criteria to evaluate the clustering results derived using different combinations of distance measures and clustering methods. The number of clusters is set to 3 as it achieved the best scores for both types of criteria in most situations. Based on the internal criteria, each clustering method achieved better scores under some CDTW bandwidths, but kernel k-means is favored in this study as it is more robust and stable against small changes in distance matrix.

From the visualizations of the clusters, we can find distinctive physical activity characteristics of each cluster in either temporality or intensity. With larger Sakoe-Chiba bandwidth in CDTW, there will be less constraint on the temporal difference between matched activities. Subsequently, the intensity difference between clusters will be more prominent than temporal difference. Moreover, the regression analysis as external criteria demonstrated that clinically meaningful differences [19] in health status indicators can be found among the four clusters. This investigation demonstrates that multiple aspects of physical activity such as intensity, duration and time may be incorporated and patterned, capturing the multiple behaviors related to physical activity as it occurs throughout a day.

In this paper, we have performed cluster analysis focusing on physical activity solely, and discovered temporal patterns and their links to health status indicators. Yet, dietary intake and physical activity may be connected in complex ways and have a cumulative influence on obesity and chronic diseases [23]. Other multi-faceted daily activities such as dietary intake may be influenced by the varying factors of physical activity behaviors over time. Their integration and the integration of further behavioral and health information to create temporal patterns may have synergistic links to health outcomes. Therefore, future studies should consider integrating additional multi-faceted behaviors.

## Data Availability

the National Health and Nutrition Examination Survey website

https://wwwn.cdc.gov/nchs/nhanes/ContinuousNhanes/Default.aspx?BeginYear=2003

